# Evaluation of Genomic Proximity Mapping (GPM) for Detecting Genomic and Chromosomal Structural Variants in Constitutional Disorders

**DOI:** 10.1101/2025.04.23.25326303

**Authors:** He Fang, Stephen M. Eacker, Yu Wu, Cate Paschal, Mary Wood, Brad Nelson, Alexander Muratov, Yajuan Liu

**Affiliations:** Department of Laboratory Medicine & Pathology, University of Washington, Seattle WA 98195; Phase Genomics, Seattle WA 98109; Department of Laboratories, Seattle Children’s Hospital, Seattle, Washington; Department of Laboratory Medicine and Pathology, University of Washington and Seattle Children’s Hospital, Seattle, WA 98195

## Abstract

Genomic structural variants (SVs) are critical contributors to genetic diversity and disease, yet their detection remains challenging with conventional cytogenetic techniques, such as karyotyping, fluorescence in situ hybridization (FISH), and chromosome microarray analysis (CMA). These methods often lack the resolution and sensitivity needed for comprehensive characterization of chromosomal aberrations. To address these limitations, we implemented genomic proximity mapping (GPM), a genome-wide chromosome conformation capture technology, in a clinical setting.

In this study, we applied GPM to a cohort of 123 patients with constitutional disorders, achieving a 100% concordance rate in detecting 411 CNVs and 39 structural rearrangements, in addition to novel findings missed by standard methods. GPM demonstrated unique advantages, such as resolving both balanced and unbalanced chromosomal rearrangements with precise (<5kb) breakpoint resolution, maintaining robust performance with challenging samples, including formalin-fixed, paraffin-embedded (FFPE) tissues, and detecting mosaicism with high sensitivity. Furthermore, GPM reliably provided detailed copy number and loss-of-heterozygosity profiles, streamlining workflows and enhancing diagnostic resolution.

GPM represents a transformative tool for genomic diagnostics, offering a high-resolution, comprehensive approach to detecting diverse genomic alterations. Its ability to address limitations of conventional cytogenetics methods positions GPM as a needed advance in the diagnosis, prognosis, and therapeutic management of genetic disorders.

## Introduction

Structural variants (SVs) are significant genomic alterations spanning at least 50 base pairs (bp) [1]. These variants are typically categorized as deletions, duplications, inversions, insertions, and translocations, encompassing both balanced and unbalanced DNA rearrangements [2, 3]. Broadly, SVs can be classified into two categories: copy-number variants (CNVs), which typically involve simple deletions and duplications that alter genomic dosage, and structural rearrangements, which reorganize DNA structure (such as inversions, insertions, and translocations) and may or may not affect copy number. SVs are usually characterized as individual events, though more intricate cases involving combinations of different SV types can also occur. For example, complex SVs have been identified in human cells, involving multiple distinct SV signatures within a single mutational event, ranging from CNV-flanked inversions to localized chromosomal shattering like chromothripsis [4, 5].

Recent studies utilizing cytogenetic and sequencing technologies have revealed that structural variation in the human genome is extensive and complex, significantly contributing to genetic diversity [6]. A typical human genome harbors thousands of germline SVs, ranging in size from kilobases to entire chromosomes. Although most SVs are benign, a subset is phenotypically relevant. Notably, SVs may account for over 25% of protein-truncating events per genome [6]. Studies have also highlighted the critical role of SVs in driving functional changes and their contributions to a variety of genetic disorders, including sporadic developmental syndromes, Mendelian diseases, complex disorders, and health-related metabolic phenotypes[6, 7]. For example, complex structural variants have been implicated in Coffin-Siris syndrome (ARID1B), hypotonia (HNRNPU), and early infantile epileptic encephalopathy with Rett-like features (CDKL5) [8].

Diagnostic testing for constitutional disorders is vital for clinical management, genetic counseling, and family planning. Chromosomal microarray (CMA) testing has been recommended as the first-tier diagnostic tool for patients with structural birth defects and developmental disabilities, superseding G-banded karyotype analysis due to its higher resolution and diagnostic yield [9]. However, CMA has technical limitations, including its inability to detect balanced rearrangements and to precisely locate duplicated DNA fragments. More recently developed and advanced sequencing-based approaches, such as whole-genome sequencing (WGS) and long-read sequencing (LRS), offer higher resolution for SV detection but have distinct limitations: WGS requires substantial sequencing depth, while LRS demands intact long DNA strands, making it unsuitable for samples with fragmented DNA, like formalin-fixed paraffin-embedded (FFPE) tissues [8, 10]. Moreover, these methods often perform poorly in repetitive regions such as centromeres and telomeres, where large-scale chromosomal rearrangements, including Robertsonian translocations, frequently occur [3, 11].

High-throughput chromosome conformation capture sequencing, known as Hi-C, forms the methodological basis of genomic proximity mapping (GPM). GPM uses Hi-C sequencing to provide a sequence-efficient approach to detecting large-scale chromosomal rearrangements [12–15]. In Hi-C, genomic loci making three-dimensional contact with each other are converted into linked read-pairs by proximity ligation of restriction-digested fixed chromatin. Large-scale chromosomal rearrangements bring together loci from different chromosomes, or that were far apart on the same chromosome, altering the Hi-C signal in predictable ways. Notably, Hi-C has been optimized for FFPE samples, allowing these methods to be applied to archived patient samples. GPM interprets alterations in Hi-C interactions to determine the linear structure of chromosomes.

Our study demonstrates that GPM is a promising tool for comprehensive SV analysis, with potential applications in enhancing the diagnosis, prognosis, and therapeutic management of genetic disorders.

## Methods

### Sample Selection and Preparation

Samples were retrospectively selected from archived clinical specimens previously tested using standard-of-care (SOC) cytogenetic methods (karyotyping, chromosomal microarray analysis (CMA), and fluorescence in situ hybridization(FISH)) in our laboratory. To ensure a representative evaluation of GPM’s diagnostic utility, cases were selected to encompass diverse specimen types, structural variants, and clinical heterogeneity. Samples were prioritized based on sample availability, DNA quality, and SOC-confirmed structural aberrations. Archived samples were stored at −80°C until processing. To minimize bias, all specimens were de-identified prior to Hi-C library preparation and analysis. Clinical metadata (e.g., sex, specimen type) and SOC results were masked during data interpretation, with concordance assessments performed post hoc.

Five lymphoblastoid cell lines with well characterized structural variants are also included to benchmark the performance of GPM. Lymphoblastoid cells lines used in this study were purchased from Coriell. Cells were cultured in RPMI 1640 with L-glutamine supplemented with 15% fetal bovine serum at 37°C in 5% CO_2_ following the provider’s protocol. A total of 500,000 cells counted by hemocytometer were used as input for the Proximo Human Hi-C kit.

### Cytogenetics and Chromosomal SNP Microarray Analysis (CMA)

Karyotyping and FISH were performed according to standard procedures (TGA manuals). CMA of genomic DNA was performed using the Illumina Infinium CytoSNP-850K BeadChip v1.1. Microarray data were visualized and analyzed using Illumina BlueFuse Multi v4.4 (Illumina, Inc., USA) and NxClinical version 10.0 (Biodiscovery, Inc., USA). All analyses and genomic coordinates reported in this manuscript are based on the human genome build GRCh38/hg38.

### High-throughput chromosome conformation capture sequencing (Hi-C; genomic proximity mapping, GPM)

Hi-C libraries were generated using the Phase Genomics Proximo Human kit v4.0 (Phase Genomics Inc., USA) following the manufacturer’s protocol. In brief, cell pellets from the peripheral blood samples previously processed by red blood cell lysis buffer (Qiagen) were crosslinked, preserving the chromatin structure within the intact nucleus. Snap-frozen chorionic villus and amniocyte samples were directly crosslinked following the Phase Genomics protocol. For FFPE tissue, chromatin was liberated from 2 x 5 µm curls by adaptive focused acoustics using a Covaris S220 instrument. Following cell lysis, chromatin was immobilized on magnetic beads and digested using restriction enzymes. The overhangs were filled in with biotinylated nucleotides and subjected to proximity ligation. Ligated junctions were purified using streptavidin beads and converted to a standard dual-indexed Illumina-compatible library.

In total, we sequenced ∼150 million pair-end reads for samples collected in our cohort. Sequencing reads were mapped to HG38 using BWA the Hi-C flags. The frequency of spatial contact is represented with heatmaps [16]. CNVs can be detected from Hi-C sequencing data using traditional sequence coverage and allele-frequency based methods, and in this study were evaluated using NxClinical version 10.0 (Biodiscovery, Inc., USA). Sequence data generated as a part of this study were also analyzed using the CytoTerra GPM cloud analysis platform (Phase Genomics, Inc., USA). Using a suite of automated variant calling algorithms, CytoTerra identified the variants described in this study, providing independent support for the manual interpretations presented here. All analyses and genomic coordinates reported in this manuscript are based on the human genome build GRCh38/hg38.

The Hi-C protocol incorporates several quality control (QC) metrics at both pre-analytical and analytical stages to ensure reliable results. Manufacturer’s recommendation for pre-analytical QC mandated a minimum input of 10 ng proximity-ligated DNA and a final library yield concentration exceeding 1 ng/μl were followed.

### Optical Genome Mapping (OGM)

Ultra-high molecular weight (UHMW) DNA was extracted from white blood cells and labeled following the manufacturer’s protocols (Bionano Genomics, USA). The fluorescently labeled DNA molecules were loaded on flow cells and imaged sequentially across nanochannels on a Saphyr instrument. A median coverage of > 100x was achieved for both samples. The proprietary OGM-specific software – Bionano Access and Solve (versions 1.6/1.7 and 3.6/3.7, respectively), were used for data processing. De novo assembly was performed using Bionano’s custom assembler software program based on the Overlap-Layout-Consensus paradigm. SVs were identified based on the alignment profiles between the de novo assembled genome maps and the Human Genome Reference Consortium GRCh38/hg38 assembly. Fractional copy number analyses were performed from alignment of molecules and labels against GRCh38/hg38.

## Results

### Study Cohort: Patients with Constitutional Disorders

Our study analyzed a cohort of 123 patients diagnosed with constitutional disorders caused by genomic structural variants, identified through standard-of-care methods. The cohort consisted of 63 males and 60 females, encompassing a diverse range of sample types collected from different developmental stages. Prenatal samples comprised 88 cases, including 49 amniotic fluid specimens, 7 chorionic villus sampling (CVS) specimens, and 32 cord blood samples. Postnatal specimens included 35 cases, comprising 27 peripheral blood samples and 8 products of conception (POC) samples. Additionally, we included 40 normal control samples, equally split between 20 males and 20 females, together with 5 cell lines with well characterized structural variants, to serve as benchmarks in the analysis.

All cases underwent SOC cytogenetic analyses, including karyotyping, FISH, and CMA. These methods collectively identified a variety of structural rearrangements and copy number alterations, which served as a benchmark for comparison with GPM results. In the study cohort of 123 patients, 411 copy number variants (CNVs) were identified, alongside 39 structural rearrangements which are distinct from simple deletions or duplications, including 26 inter-chromosomal and 13 intra-chromosomal events. As a positive control, all known structural variants were confirmed in five cell lines, while the 40 normal control samples showed no clinically significant structural variants.

Of the 163 samples selected (including 123 samples with constitutional disorders and 40 normal control samples), 5 failed QC criteria, resulting in a pass rate of 97%. These failures were attributed to insufficient library yield or degraded DNA quality due to prolonged storage, which prevented successful library preparation or sequencing. The failed samples included 1 amniotic fluid, 2 cord blood, and 2 peripheral blood specimens, emphasizing that archival sample quality rather than sample type primarily affects success rates.

### Identification of Balanced and Unbalanced Structural Rearrangements Using GPM

Chromosomal structural rearrangements are relatively prevalent chromosomal abnormalities in the general population, occurring in up to 1 in 500 live births [17]. Balanced structural rearrangements manifest in adults experiencing reproductive challenges, such as subfertility or recurrent pregnancy loss [18]. Conversely, unbalanced structural rearrangements are frequently observed in children exhibiting congenital abnormalities, developmental delays, or intellectual disabilities. In clinical laboratories, the standard method for detecting chromosomal structural rearrangement involves karyotyping, supplemented by FISH when a specific translocation is suspected. However, these techniques are constrained by limited resolution capabilities, or in the case of FISH, the need for variant-specific probes. Therefore, assessing the efficacy of Hi-C in detecting both balanced and unbalanced structural rearrangements within clinical settings was an important focus of our technical evaluation.

In this study, structural rearrangements identified by GPM are annotated by CytoTerra and further manually curated using Hi-C heatmaps. The Hi-C heatmap represents pairwise sequence interaction frequencies across the genome, where sequences in close physical proximity on a chromosome interact more frequently than those further apart. Below briefly describes how to interpret the Hi-C data presented in this study.

Structural rearrangements appear on Hi-C heatmaps as deviations from the expected pattern of sequence interactions. For example, a translocation manifests as an excess of interchromosomal interactions between the involved chromosomes. The breakpoint can be identified as the region with the highest levels of interchromosomal interactions, with signal intensity decaying according to a power law function as it extends from the breakpoint [15]. Intrachromosomal SVs follow similar principles, with deviations observed in expected intrachromosomal sequence interactions. Breakpoints are identified by regions of unexpected pairwise interactions, again following a power law decay as the signal extends outward. Representative Hi-C heatmaps in Figure 1A illustrate these characteristic disruption patterns identified by GPM, representing reciprocal translocation, nonreciprocal translocation, insertion, inversion, deletion and duplication, respectively from top to bottom, left to right.

**Figure 1.**
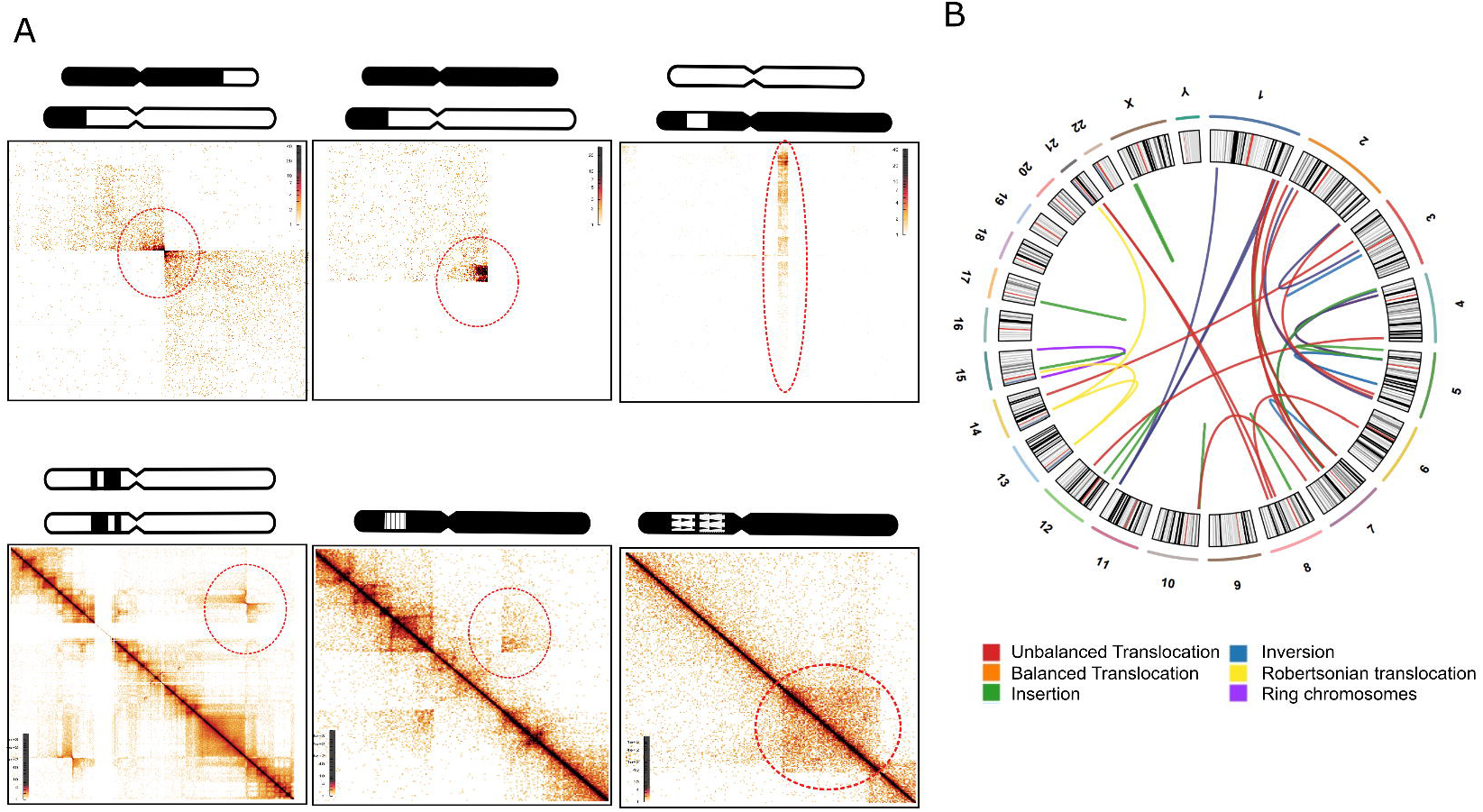
Structural Variants Detected by GPM. (A) Hi-C heatmaps illustrating disruption patterns associated with different types of structural variants. The six heatmaps (arranged from left to right, top to bottom) depict distinct patterns of balanced translocation, unbalanced translocation, insertion, inversion, deletion, and duplication. (B) Circos plot showing all structural rearrangements identified in our study cohort, excluding copy number variants. The plot includes balanced and unbalanced translocations, insertions, inversions, Robertsonian translocations, and ring chromosomes.

Within our study cohort, we identified a collective 39 structural rearrangements events in a total of 14 cases. These encompassed 26 inter-chromosomal and 13 intra-chromosomal rearrangements (Figure 1B, Supplementary Table 1). The spectrum of translocation types includes reciprocal translocations (n=8), nonreciprocal translocations (n=13), Robertsonian translocations (n=3), inversions (n=4), insertions (n=10), and one instance of ring chromosome (n=1). The size of fragments involved in these structural rearrangements spanning from **719 kb** to the whole chromosome arm. In all these 39 translocation events, concordance of GPM results and SOC results was observed in 100% of cases (Figure 1B). Furthermore, GPM not only matched the SOC results but also refined the breakpoint resolution. Additionally, GPM identified novel findings missed by SOC methods, demonstrating its capacity to uncover structural variants that traditional techniques failed to detect (see details below). This precision was achieved with a sequencing depth of approximately 50 million high quality pair-end reads.

A. Identification of submicroscopic structural rearrangements with high precision Hi-C identified both reciprocal and nonreciprocal translocations in the cohort, achieving a resolution of 10 kb to 100 kb depending on sequencing coverage. A notable example of this capability was the precise characterization of a familial t(2;5) translocation, as shown in Figure 2A and 2B (see [15] for details). This case involved three family members with a history of t(2;5), where a ∼3 Mb terminal fragment on chromosome 2 and a ∼4 Mb terminal fragment on chromosome 5 were translocated. Traditional chromosome analysis failed to detect this translocation due to its small size and terminal location. Consequently, the family underwent additional tests, including CMA and WGS, which revealed reciprocal unbalanced copy number changes in two symptomatic members. These results suggested a balanced translocation in the asymptomatic parent. GPM analysis was then conducted on all three family members to confirm and further delineate the accurate translocation breakpoints. GPM successfully resolved the translocation breakpoints at the gene level. In the Hi-C heatmap (Figure 2), the asymptomatic carrier exhibited two high-intensity “hot corners” corresponding to reciprocal breakpoints between chromosomes 2 and 5 in Figure 2A, whereas symptomatic family members showed a single hot corner in Figure 2B, indicative of their unbalanced rearrangements. The patient harboring only the derivative chromosome 5 displayed increased interaction between the telomeric region of chromosome 2 and a large segment of chromosome 5, while the patient with only the derivative chromosome 2 showed enhanced contact between the telomeric region of chromosome 5 and a large segment of chromosome 2. The breakpoint on chromosome 2 is located between chr2:238,950,000 and chr2:238,960,000, with the critical gene *TWIST2* nearby remaining intact and unaffected by the rearrangement, suggesting its function is likely preserved (Figure 2A and 2B left). Similarly, the breakpoint on chromosome 5 is located between chr5:174,950,000 and chr5:174,960,000, with no critical gene identified in this region (Figure 2A and 2B right). These results underscore the ability of GPM to accurately characterize submicroscopic chromosomal rearrangements, offering insights that were unattainable with conventional cytogenetic methods. Moreover, GPM eliminated the need for sequential cytogenetic and microarray testing in familial cases with possible inherited rearrangements. In one case in our cohort, karyotype and CMA of a POC sample from a terminated pregnancy revealed an unbalanced derivative of chromosomes 4 and 5, prompting follow-up analyses that confirmed the mother’s balanced rearrangements involving a t(4;5)(p15.2;p13.3) and an inv(5)(p13.3q23). In a subsequent pregnancy, chorionic villus sampling (CVS) was analyzed first by conventional karyotype—which identified the translocation and the pericentric inversion—and then by CMA, which suggested the inheritance of balanced status. However, these methods were insufficient, where the translocation breakpoints could only be inferred, not confirmed, by microarray and the inversion breakpoints were detectable solely by karyotype with limited resolution. GPM resolved a complex series of rearrangements in a single assay with gene level resolution, as shown in **Figure 2C**. Block ‘a’, as circled in blue square, in the Hi-C heatmap shows increased interaction between a portion of the short arm of chromosome 5 and the most part of chromosome 4, while Block ‘b’, also circled in blue square, reveals increased interaction between a portion of the short arm of chromosome 4 and the most part of chromosome 5. Additionally, Block ‘c’, along with the hot corner indicated by the arrow within Block ‘b’, confirms the presence of a pericentric inversion on chromosome 5. These findings align with the derivative chromosomes 4 and 5 detected by karyotyping (Figure 2C bottom). Additionally, it mapped the translocation breakpoints to 4p15.2 (chr4:23,540,395) and 5p13.3 (chr5:31,385,339) and delineated the inversion boundaries at 5p13.2 (chr5: 36,367,799) and 5q23.2 (chr5:125,089,272) (Supplementary Figure 1). This comprehensive characterization, achieved without sequential testing, highlights GPM’s ability to resolve complex rearrangements and provides critical breakpoint resolution for recurrence risk counseling. Moreover, this case demonstrates that GPM excels at resolving subtle structural rearrangements even in complex scenarios. With a zoomed in view, a minor change was observed within block ‘a’, where the overall pattern indicated a translocation between the short arms of chromosome 4 and chromosome 5 (Figure 2C). A closer examination of the hotspot revealed that the region of highest intensity is located within the block rather than at the expected corner of the square, alongside with a stripe of absence of interactions, as indicated by the red circle in the upper panel of Figure 2D. Moreover, further analysis of chromosome 5 uncovered additional alterations beyond the large inversion, including an intensified stripe, the absence of a stripe within the inversion block, and an additional inversion "bowtie" pattern near the diagonal, as indicated by the red circles in the lower panel of Figure 2D. These observations led us to conclude that chromosome 5 not only harbors a large inversion between chr5:36,367,799 and chr5:125,089,272, but also an insertion of a fragment from chr5:34,249,800 to chr5:34,979,800 into the region near chr5:36,367,799 (magenta line in Figure 2E), and a smaller inversion between chr5:31,379,400 and chr5:36,336,300. The translocation between chromosomes 4 and 5 features a breakpoint on chromosome 5 at position 36,336,300, situated just above the inserted segment. The complex rearrangements resolved by GPM is shown by the ideogram in Figure 2E. Due to the presence of the small insertion on chromosome 5 which is not involved in the translocation between chromosomes 4 and 5 (5p13.2, chr5:34,200,000-35,000,000, Supplementary Figure 2A), in the unbalanced fetus terminated—who carries two normal copies of chromosome 4, one normal copy of chromosome 5, and one derivative chromosome 5—the inserted fragment on the derivative chromosome is effectively retained in two copies. This finding aligns with our array results, which revealed two discontinuous deletions on chromosome 5, and confirms that the dosage-sensitive gene DNAJC21 remains unaffected (Supplementary Figure 2B). Overall, this case illustrates how GPM can detect cryptic, complex rearrangements that might be missed by traditional cytogenetic methods. In our cohort, the smallest fragment involved in a structural rearrangement we identified was an insertion of a 719 kb fragment from chromosome 7 to chromosome 1 (Supplementary Figure 1C). The Hi-C analysis pinpointed the precise breakpoints of this insertion with gene-level resolution, revealing two breakpoints on chromosome 7 and two on chromosome 1. This insertion is depicted in Supplementary Figure 2C as a long string of high-intensity interactions formed between a small region of chromosome 1 to the almost entire chromosome 7. The intensity distribution is uneven, with the highest intensity observed at the breakpoint on chromosome 7, clearly highlighting the precise coordinates of the breakpoints. This insertion represents a novel finding that was missed by conventional cytogenetic methods due to its small size but was subsequently verified using other technologies, including OGM (data not shown). Notably, this insertion is part of a complex chromosomal rearrangement case with more than 10 breakpoints, where Hi-C proved invaluable in unraveling the intricate structural details (in press for GIMO). This comprehensive analysis encompassed a spectrum of genetic alterations, including insertions, inversions, as well as inter- and intra-chromosomal translocations.
B. Identification of Robertsonian translocations and whole arm translocations GPM also demonstrated distinct advantages in the identification of Robertsonian and whole-arm translocations. In our study cohort, we identified eight cases of trisomy, two of which were associated with Robertsonian translocations. We also included a cell line derived from a balanced carrier of a t(13;15) Robertsonian translocation [19]. Our findings confirm that GPM analysis effectively distinguishes free trisomies from those involving Robertsonian translocations. To illustrate this, we present Hi-C results from two POC cases, all of which also underwent testing using SOC methods (Figure 3). Case A was confirmed to have trisomy 13 by FISH and karyotyping, revealing three independent copies of chromosome 13. This was reflected by a global increase in interaction frequency for chromosome 13 by GPM, seen as a darker stripe across the Hi-C heatmap. Case B involved trisomy 14 due to a Robertsonian t(13;14) translocation. Hi-C heatmap revealed both a darker stripe for chromosome 14 across the whole genome, indicating a copy gain, and an increased interaction between chromosomes 13 and 14. Notably, a strong interaction pattern was observed between the p-terminal (p-ter) regions of chromosomes 13 and 14, forming a distinct darkened corner in the contact map—an interaction signature characteristic of Robertsonian translocations. The copy number changes associated with these trisomies can also be demonstrated by the increased sequencing coverage, as indicated by the coverage track in the top panel of Figure 3A and 3B. Case C was a cell line derived from a balanced carrier of a Robertsonian t(13;15) translocation. Unlike the trisomic cases, Hi-C did not show a copy gain for any chromosome. However, a distinctive interaction pattern between the p-ter regions of chromosomes 13 and 15 was clearly visible, again forming a darkened corner, consistent with the p-ter-to-p-ter fusion characteristic of Robertsonian translocations. Additionally, our study highlights the robustness of the Hi-C methodology, especially its effectiveness in analyzing POC samples that are commonly formalin-fixed and paraffin-embedded for storage. In our study, we identified 5 translocation events with 100% concordance in the FFPE samples. To illustrate this, we present Hi-C data from an FFPE sample (case D) from an external patient (Figure 3D). An initial karyotype performed externally revealed an abnormal chromosome 22 with additional, uncertain material on its short arm (add(22)(p10)); however, because the sample was fixed for storage and shipment, a repeat karyotype was not possible upon receipt. The GPM analysis with the FFPE sample identified an unbalanced whole-arm translocation between the short arm of chromosome 4 and chromosome 22 [der(22)t(4;22)(p10;p10)], a finding confirmed by SNP array analysis, which showed a copy gain of the entire short arm of chromosome 4 (Supplementary Figure 2D). This identification not only clarifies the origin of the aberrant material on chromosome 22 but also suggests a potential recurrent risk that warrants follow-up for the patient’s mother.

**Figure 2.**
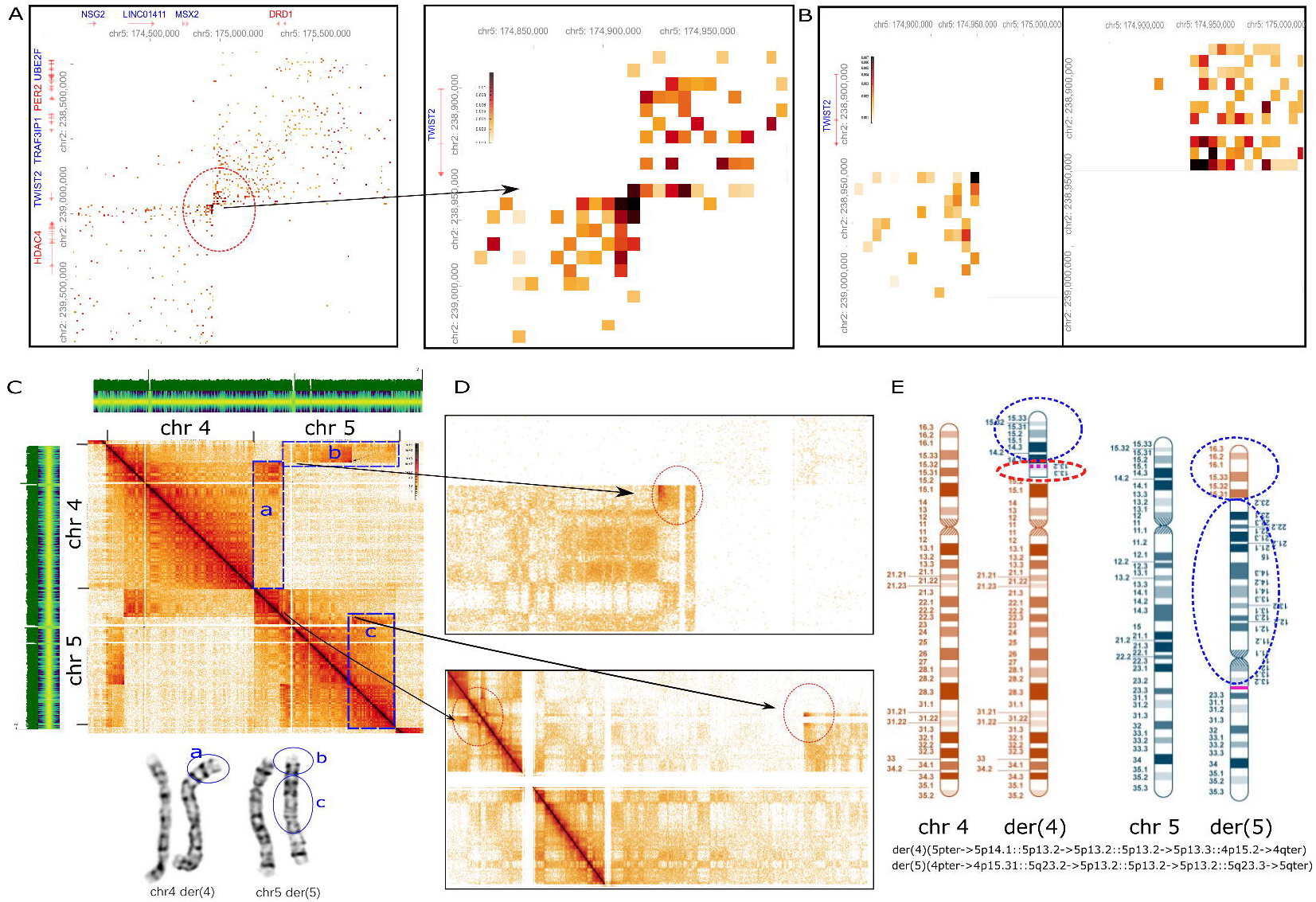
Balanced and Unbalanced Translocations Detected by GPM. (A and B) Detection of balanced and unbalanced t(2;5) translocations within a family. (A) shows a balanced t(2;5) translocation in an asymptomatic family member, with a zoomed-in view highlighting the breakpoint at gene-level resolution. (B) shows unbalanced t(2;5) translocations in two symptomatic family members, with a corresponding zoomed-in view of the breakpoint. The critical genes MSX2 and TWIST2 are not directly involved, as the translocation breakpoints (hot corners) are located outside these gene regions. (C and D) A complex chromosomal translocation identified in a balanced translocation carrier. (C) shows an overall view of the rearrangements between chromosome 4 and chromosome 5. The lower panel shows the derivative chromosome 4 and 5 analyzed by karyotype. a, b, and c annotate the regions involved in the rearrangements. (D) shows the zoomed-in view of the sub-microscopic rearrangements on chromosome 5, including a small insertion and a small inversion in addition to the large inversion on chromosome 5. (E) shows the ideogram illustrating the complex rearrangement. The regions within the blue circle are rearrangements detected by chromosomal analysis. The regions within the red circle are novel findings detected by GPM.

**Figure 3.**
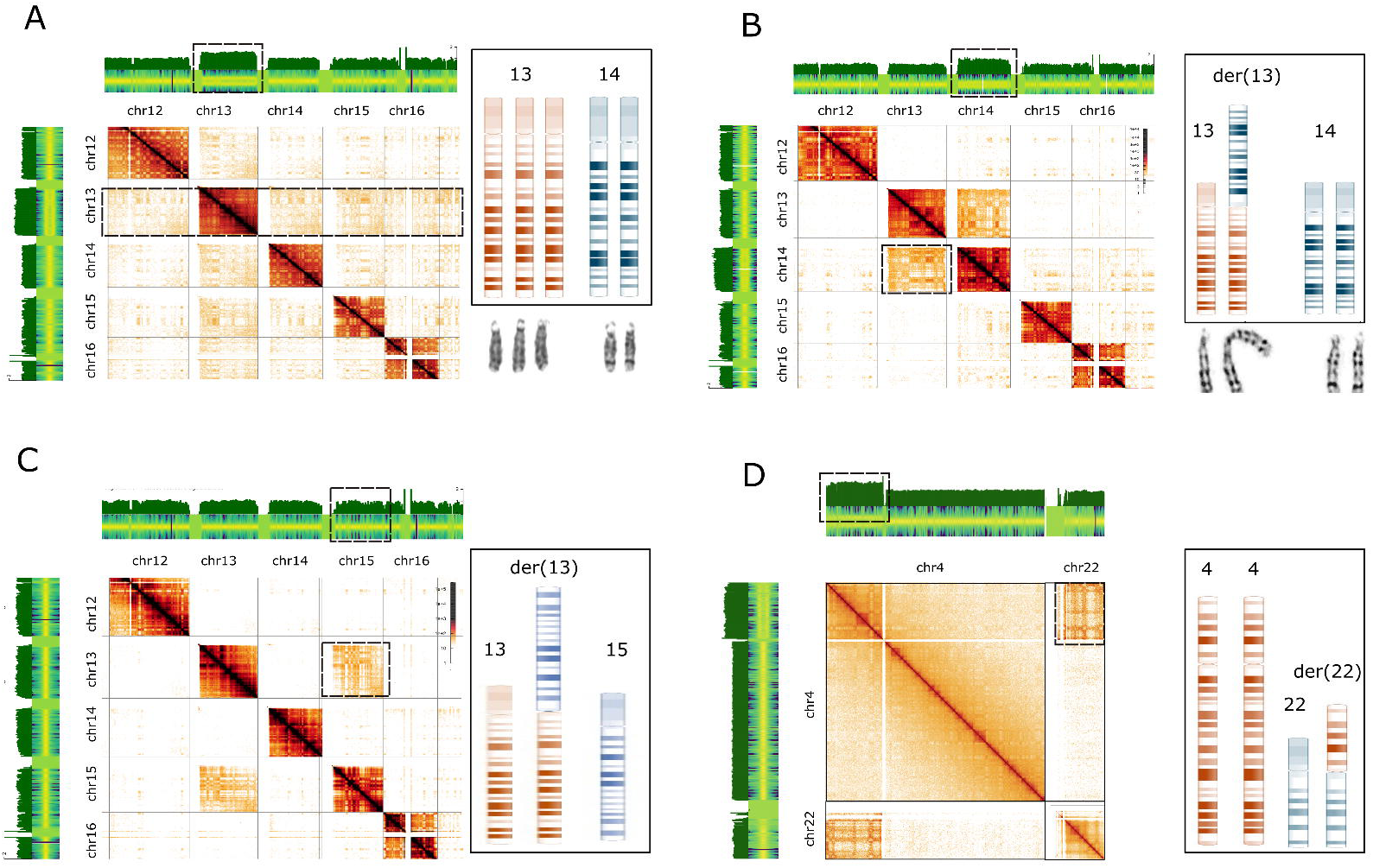
Robertsonian and whole-arm translocations detected by GPM. (A) Detection of free trisomy 13 by GPM and chromosomal analysis. The copy gain of chromosome 13 is evident from increased sequencing coverage and heightened intensity of chromosome 13 across the genome in the Hi-C heatmap (black box). The corresponding ideogram is shown on the right. (B) Detection of an unbalanced Robertsonian translocation, der(13;14). The copy gain of chromosome 13 is indicated by increased sequencing coverage and heightened intensity in the Hi-C heatmap. The presence of the Robertsonian chromosome is confirmed by increased interaction intensity between chromosomes 13 and 14 in the Hi-C heatmap. The ideogram is shown on the right. (C) Detection of a balanced Robertsonian translocation, der(13;15). The Robertsonian chromosome is identified by increased interaction intensity between chromosomes 13 and 15 in the Hi-C heatmap. The ideogram is shown on the right. (D) Detection of a whole-arm translocation between chromosomes 4 and 22. The copy gain of 4p is evident from increased sequencing coverage and heightened intensity of chromosome 4p in the Hi-C heatmap. The translocation is indicated by increased interaction intensity between chromosome 4p and chromosome 22 in the Hi-C heatmap. The ideogram is shown on the right.

### Identification of Copy Number Alterations with GPM

We also evaluated the performance of GPM in identifying copy number alterations with our study cohort. In our analysis, we identified a total 411 CNV events—235 deletions and 176 duplications. One example is a ∼1.8 Mb deletion on chromosome 6, identified in a cord blood sample from a fetus whose father carries the same deletion (Figure 4A). This region encompasses approximately 27 genes, including the dosage-sensitive *TAB2* gene, which is associated with a spectrum of phenotypes such as growth restriction and congenital heart abnormalities [20]. The deletion was initially detected by the SNP array, as shown in the top panel of Figure 4A. GPM confirmed this deletion through multiple signals: reduced sequencing coverage (log 2 ratio (logR), top track), loss of heterozygosity in the B-allele frequency track (BAF, middle track), and a distinct hot-corner pattern in the Hi-C heatmap (Figure 4A, middle). The sequencing data generated by GPM can also be uploaded and analyzed using the same software used for SNP array analysis, NxClinical in this study. The data includes a sequencing coverage track (black), a logR track derived from coverage normalized to whole-genome sequencing depth, and a BAF frequency track. The deletion is clearly visible as a reduction in sequencing coverage and logR ratio, along with a loss of heterozygosity in the BAF track (Figure 4A, bottom). Similarly, a ∼8.6 Mb duplication on chromosome 13 was identified in a blood sample from a pregnant woman (Figure 4B). This duplication was first flagged by an abnormal cell-free DNA screening result and later confirmed by SNP array (Figure 4B, top). GPM analysis revealed this duplication through increased sequencing coverage (logR, top track), a split in the B-allele frequency track, and the appearance of an additional high-intensity square in the heatmap, further supporting the structural alteration (Figure 4B, middle and bottom).

**Figure 4.**
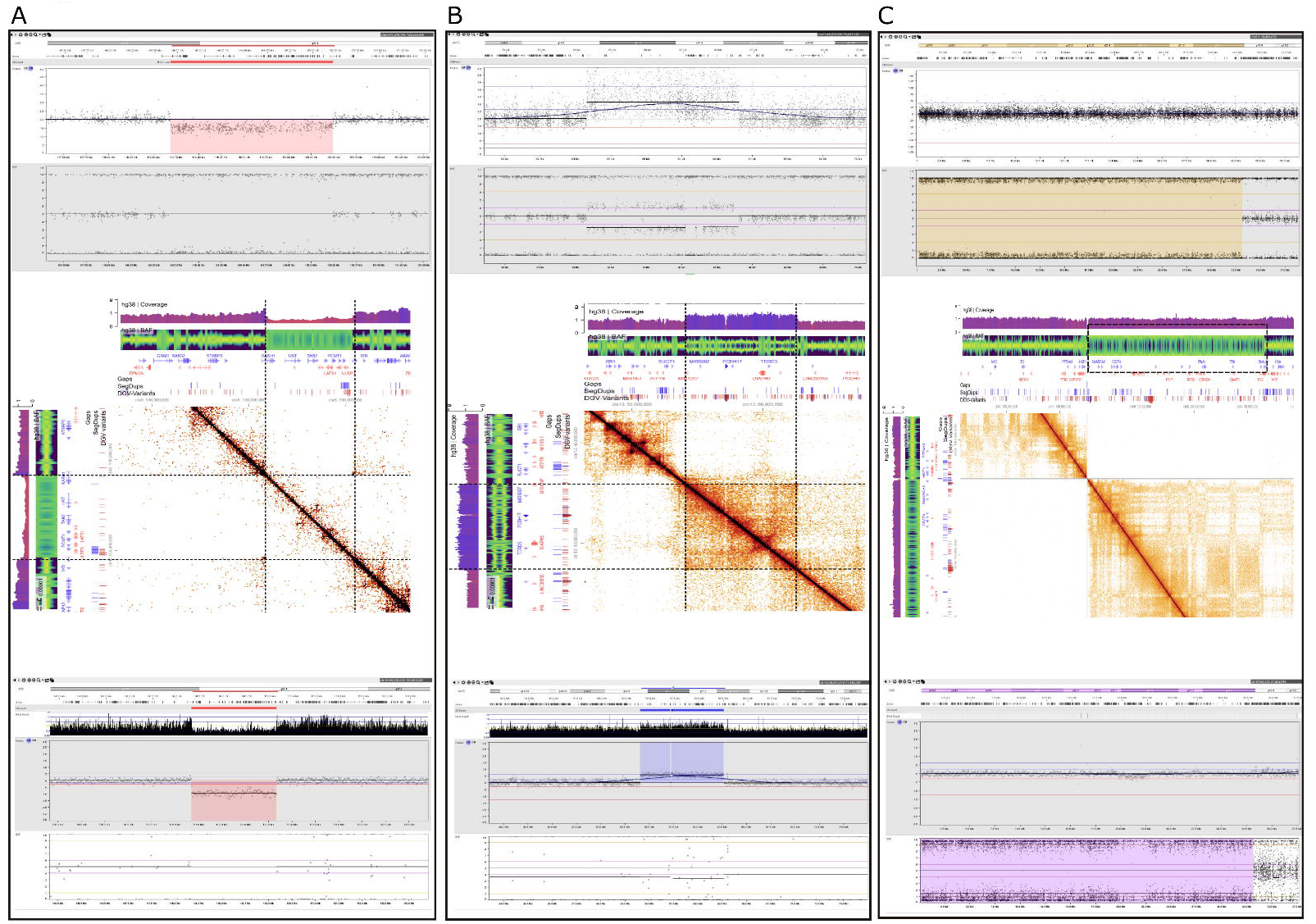
CNV detection by SNP array and GPM is illustrated for three cases—a deletion (A), a duplication (B), and an absence of heterozygosity (C). For each case, the top panel displays the SNP array result with logR and BAF tracks; the middle panel shows the corresponding Hi-C heatmap from GPM, which includes sequencing coverage, MAF, and contact intensity tracks; and the bottom panel presents the GPM data visualized using NxClinical software with sequencing depth, logR, and BAF tracks.

The accuracy of GPM in detecting CNVs depends on factors such as sequencing depth, the size of the CNV region, and the nature of the abnormality. To define the depth requirement for reliable detection across varying CNV sizes, we systematically evaluated the performance of GPM at different sequencing depths. Without applying the sequencing depth cutoff, GPM identified 217 out of 235 deletion events, corresponding to a detection rate of 92%. The 18 deletions not detected were primarily characterized by sizes smaller than 25 kb or sequencing depths below 60 million reads (Figure 5A). For samples with sequencing depths exceeding 60 million reads, our method achieved a detection rate of 94% for deletions larger than 10 kb (64/68 events) and 100% for deletions exceeding 25 kb (58/58 events). Notably, the smallest deletion detected was a 7.8 kb deletion within the Hbb gene region. This sample is from a patient known to be a thalassemia carrier. The presence and size of the deletion were confirmed by PCR analysis performed by an external laboratory (data not shown). This deletion was also seen but not called by CMA. Similarly, for duplications larger than 10 kb, the sensitivity was 97% (57/59 events), and for duplications over 25 kb, sensitivity reached 100% (52/52 events) (Figure 5B). A unique aspect of our study was the inclusion of variants of uncertain significance, likely pathogenic and pathogenic variants to enable a comprehensive evaluation. When focusing specifically on likely pathogenic and pathogenic CNVs, the GPM method demonstrated a 100% detection rate for both deletions (48/48) and duplications (37/37), further emphasizing its reliability in identifying clinically relevant CNVs.

**Figure 5.**
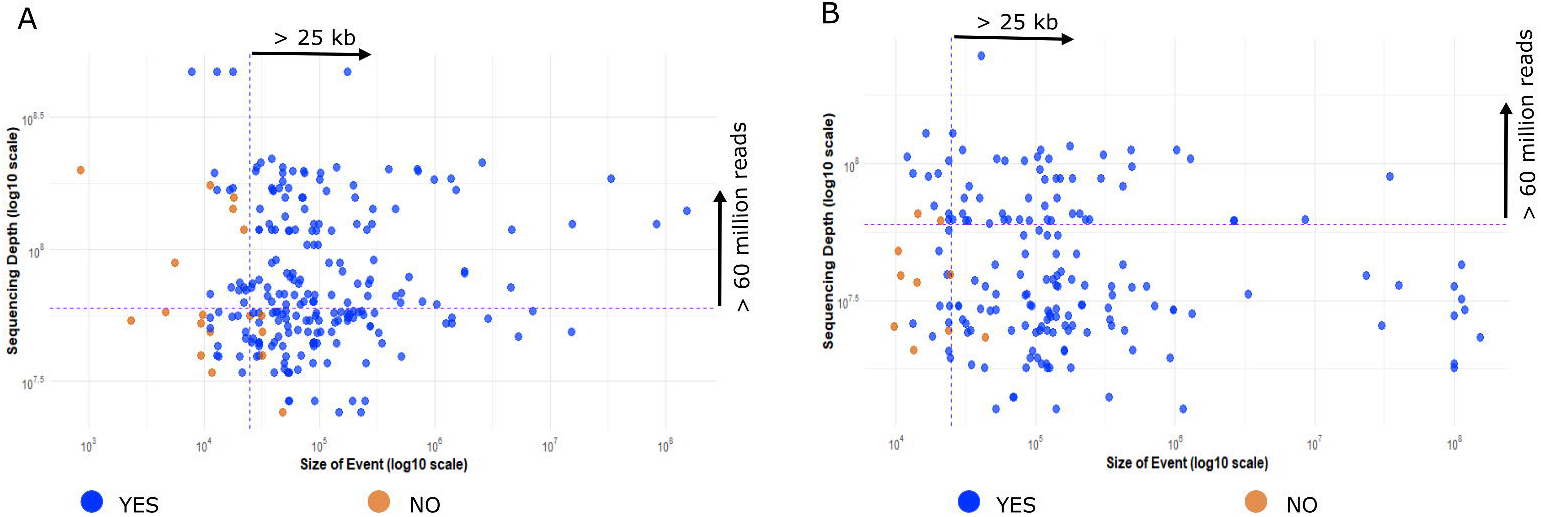
Concordance of CNV Detection by GPM and SNP Array Across CNV Sizes and Sequencing Depths. Scatter plots illustrate the detection concordance for CNVs of varying sizes and sequencing depths using both GPM and SNP array. (A) shows deletion events and (B) shows duplication events. Blue dots represent events detected by both assays, whereas orange dots indicate events missed by GPM but captured by the SNP array.

Beyond CNV detection, GPM demonstrated robust performance in identifying areas of absence of heterozygosity (AOH), comparable to other NGS-based approaches. One example is a ∼30 Mb AOH region on chromosome 9, initially detected by array analysis (Figure 4C, top panel). Since GPM also incorporates SNP information, this AOH was clearly visualized in the BAF track (Figure 4C). As expected, no changes were observed in sequencing coverage or the Hi-C heatmap, consistent with the absence of a copy number alteration. We also systematically evaluated the performance of GPM at different sequencing depths. In our study cohort, in samples with read depths exceeding 60 million, GPM detected 47 out of 51 AOH events larger than 40 kb. At higher read depths of 130 million, the method achieved 100% concordance, successfully identifying all 34 AOH events larger than 40 kb. These findings establish Hi-C as a reliable tool for AOH detection, with sensitivity closely tied to sequencing depth. However, AOH detection requires greater coverage than CNV detection; specifically, 130 million high-quality reads are needed to reliably detect AOH events larger than 40 kb.

### Determine the location and orientation of fragments within duplications

GPM can detect CNVs in part through changes in Hi-C heatmap (or as the pairwise sequence interaction frequency), enabling the ability to pinpoint the genomic location and orientation of duplicated fragments. One particular case in our cohort was a female patient with developmental delay. The sample was initially examined using array analysis, revealing the presence of two duplications, chrX:6,530,710-6,941,810 and chrX:7,630,901-8,167,603, located adjacently on the X chromosome (Figure 6A). To facilitate interpretation, we designated breakpoints from pter to qter as a (chrX:6,530,710), b (chrX:6,941,810), c (chrX:7,630,901), and d (chrX: 8,167,603). The clinical significance of both duplications are uncertain per ACMG guideline. However, the CMA results left uncertainty regarding whether these duplications arose from two separate duplication events, occurring between a and b, and c and d, respectively, or if more complex rearrangements were involved. Accurately characterizing such rearrangements is crucial for variant classification, understanding their implications for the patient, and clarifying recurrent risk.

**Figure 6.**
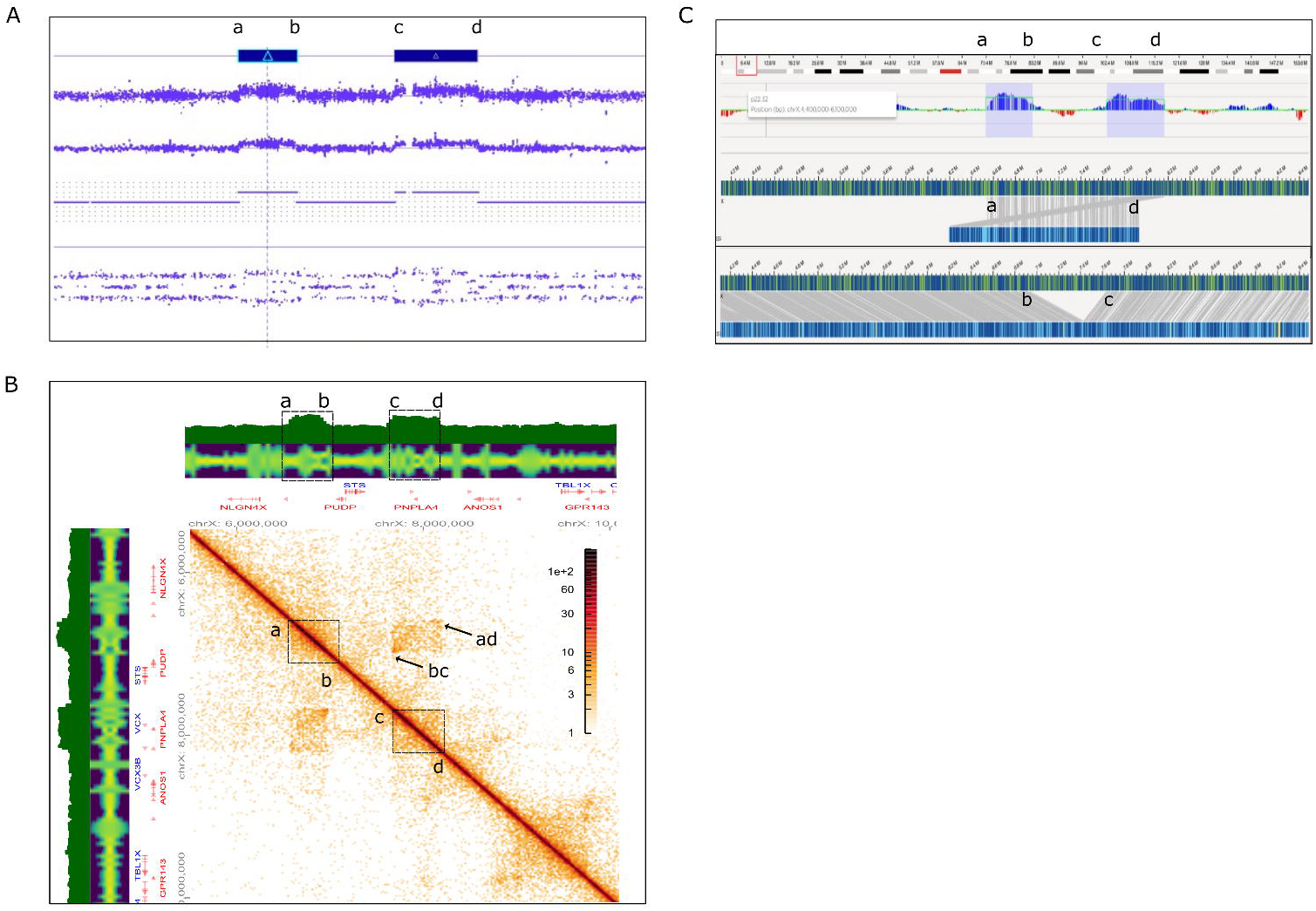
Detection of duplications associated with complex rearrangements by GPM and OGM. (A) The microarray features a top panel with logR values and a bottom panel with the B-allele frequency track, clearly identifying two duplications. (B) GPM detection of these duplications is shown by a green sequencing coverage track with increased coverage at the duplicated regions, accompanied by a heatmap that reveals elevated interaction intensities between regions a–b and c–d (outlined by square boxes). Additional interactions between a–d and b–c as indicated by arrows. (C) OGM detection of these duplications is shown with the top sequencing coverage track highlighting the gains and the lower panel displaying molecular configurations that connect regions a–d and b–c.

To address these uncertainties, we conducted GPM analysis on the sample (Figure 6B). Our results confirmed the detection of two copy number gains. These two copy gains are evident by copy number variation analysis, which shows increased sequencing coverage in the green track, as the other NGS-based CNV detection methods. Additionally, the Hi-C heatmap highlights two distinct blocks with elevated interaction intensity, marked by black squares, aligning with the identified copy gains. Moreover, the heatmap uncovered interactions between regions a and d, as well as between b and c, as the hot corners pointed by arrows, suggesting the presence of a more complex rearrangement. The presence of hot corners between regions a and d, and between b and c, suggests that the observed copy number variation results from a larger duplication spanning a–d, along with a smaller deletion between b and c (Figure 6B). These duplications are also confirmed by OGM (Figure 6C). Copy number variation analysis reveals increased coverage between a and b, as well as between c and d, highlighting the two copy gains. Additionally, the molecule configuration shows a large duplication from a to d, indicated by the connection between them, and a smaller deletion from b to c, reflected by the connection between c and d.

Detection of duplication is relatively common in practice and it prompts several questions, particularly regarding the orientation of the duplications, which could impact interpretation of genes near the breakpoints. The detection of a junction between regions a and d by both GPM and OGM suggests a tandem duplication. To further investigate, we simulated GPM data for various duplication configurations and compared the resulting heatmaps to that of our case [21]. Simulations included two separate small duplications—either tandem or inverted—and a tandem duplication containing an internal deletion. The heatmap corresponding to the tandem duplication with deletion showed two squares with distinct corner signals, a pattern not observed in the scenarios involving separate or inverted duplications (Supplementary Figure 3). Our case data closely matched the simulated pattern for a tandem duplication with an internal deletion, supporting the conclusion that the duplication is tandem in nature.

### Limit of Detection of GPM

To assess the limit of detection (LOD) for GPM, we first performed a comprehensive analysis of the sequencing coverage required to detect structural rearrangements and CNVs. For structural rearrangements, we carried out an *in silico* dilution experiment using a case with complex, balanced rearrangements involving four chromosomes and 13 breakpoints. The original sample, sequenced to yield approximately 100 million reads (with about 60 million high-quality reads, or ∼10× coverage), was down sampled to 50% and 25% of its original depth. We then evaluated how reduced coverage affected the detection of a representative abnormality as well as all SV events—both inter- and intra-chromosomal rearrangements such as insertions, inversions, and translocations. Representative Hi-C heatmaps (Figure 7A) illustrate that an interchromosomal 4.7 Mb translocation (blue circle on left) remains clearly visible even at 1× coverage, while an intrachromosomal 0.8 Mb insertion (blue circle on right) is detectable at 5× coverage but becomes challenging to identify at 1×. A comprehensive comparison of all detected events is provided in the accompanying table (Table 1). For CNV detection, we already showed that a minimum of 60 million reads is needed to detect CNVs larger than 25kb. Here we further conducted a similar in silico dilution experiment using a case with a 2.2 Mb deletion originally sequenced to yield approximately 300 million reads (∼30× coverage). The reads were downsampled to 50%, 25%, and 10% of the original dataset, and we then evaluated how reduced coverage affected the detection of the 2.2 Mb deletion. Representative logR tracks (Figure 7B) demonstrate that the deletion remains clearly visible even with 30 million reads (at 3× coverage), although smaller CNVs, duplications, and regions of absence of heterozygosity (LOH) are more challenging to detect and typically require higher sequencing depths (often exceeding 60 million reads) for consistent identification (as showed above and in Figure 6).

**Figure 7.**
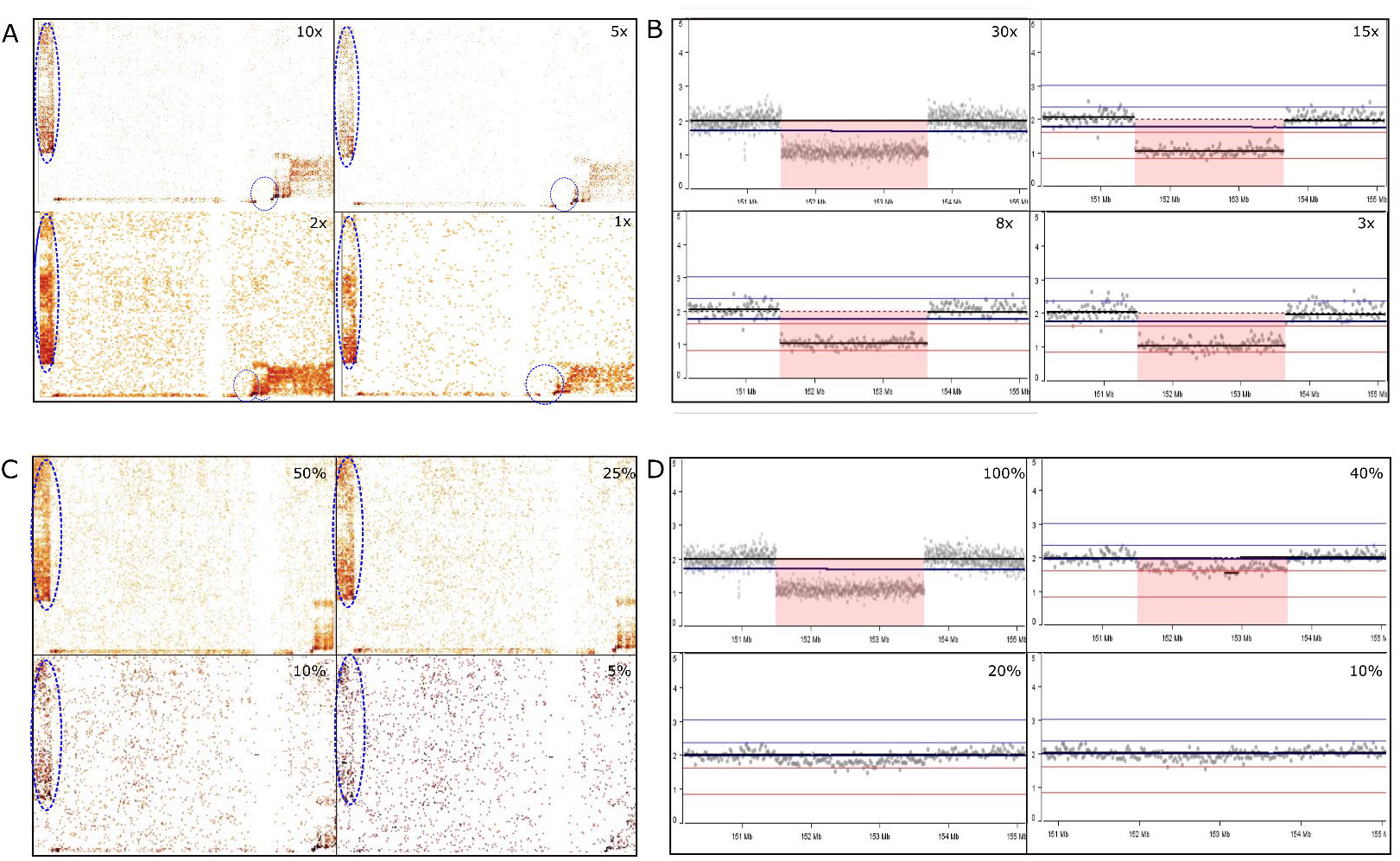
Evaluation of the detection limit of GPM for structural rearrangements and copy number variants (CNVs). (A and C) illustrate the effects of sequencing depth and mosaicism level on the detection of structural rearrangements using Hi-C heatmaps, respectively. Two rearrangements are shown: a 4.7 Mb interchromosomal translocation (circled by a large blue circle) and a 0.8 Mb intrachromosomal insertion (circled by a small blue circle). The large translocation is clearly detectable at sequencing depths of 10×, 5×, and 2×, and at mosaicism levels of 50%, 25%, and 10%. In contrast, the small insertion is detectable only at 10× and 5× coverage, and at mosaicism levels of 50% and 25%. (B and D) present the analysis for CNVs using logR values, focusing on a 2.2 Mb deletion. This deletion is observable at sequencing depths of 30×, 15×, 8×, and 3×, and at the mosaicism level is 40%, but is not detected at mosaicism levels of 20% or 10%.

We also evaluated the level of mosaicism at which GPM can detect structural rearrangements and CNVs. Given the rarity of mosaicism in constitutional disorders, we simulated varying levels by creating in silico mixtures of normal and abnormal samples. For structural rearrangements, we used the same complex rearrangement case and mixed it with a normal sample to simulate mosaicism levels of 50%, 25%, 10%, and 5%. Analysis of representative Hi-C heatmaps (Figure 7C) showed that an interchromosomal 4.7 Mb translocation is clearly visible at 50% and 25% mosaicism but becomes challenging at 10%, whereas an 86 Mb translocation remains detectable at 10% mosaicism but is difficult to identify at 5% (Figure 7C). Similarly, for CNV detection, we simulated mosaicism in the case with the ∼2.2 Mb deletion by mixing it with a normal sample to achieve mosaic levels of 40%, 20%, and 10% (Figure 7D). Representative logR tracks reveal that the deletion is clearly visible at 40% mosaicism, still discernible at 20%, but becomes undetectable at 10%. For large regions of LOH, GPM reliably detects mosaicism at 60%, although the B-allele frequency (BAF) signal becomes less distinct at lower mosaic levels (Supplementary Figure 4). Overall, these experiments demonstrate that the sensitivity of GPM for detecting both structural rearrangements and CNVs—and their mosaic forms— is strongly influenced by sequencing coverage, with higher coverage enabling improved detection of low-level mosaic events.

## Discussion

Our study demonstrates that the GPM is uniquely suited for detecting and resolving structural rearrangements by capturing three-dimensional chromatin interactions. Unlike traditional linear sequencing approaches that treat DNA as a one-dimensional sequence, Hi-C mapping within GPM reveals spatial proximities between genomic loci (Figure 8). This three-dimensional information enables GPM to detect abnormal contact patterns in chromatin interaction matrices, even when reads are not directly mapped to breakpoints.

**Figure 8.**
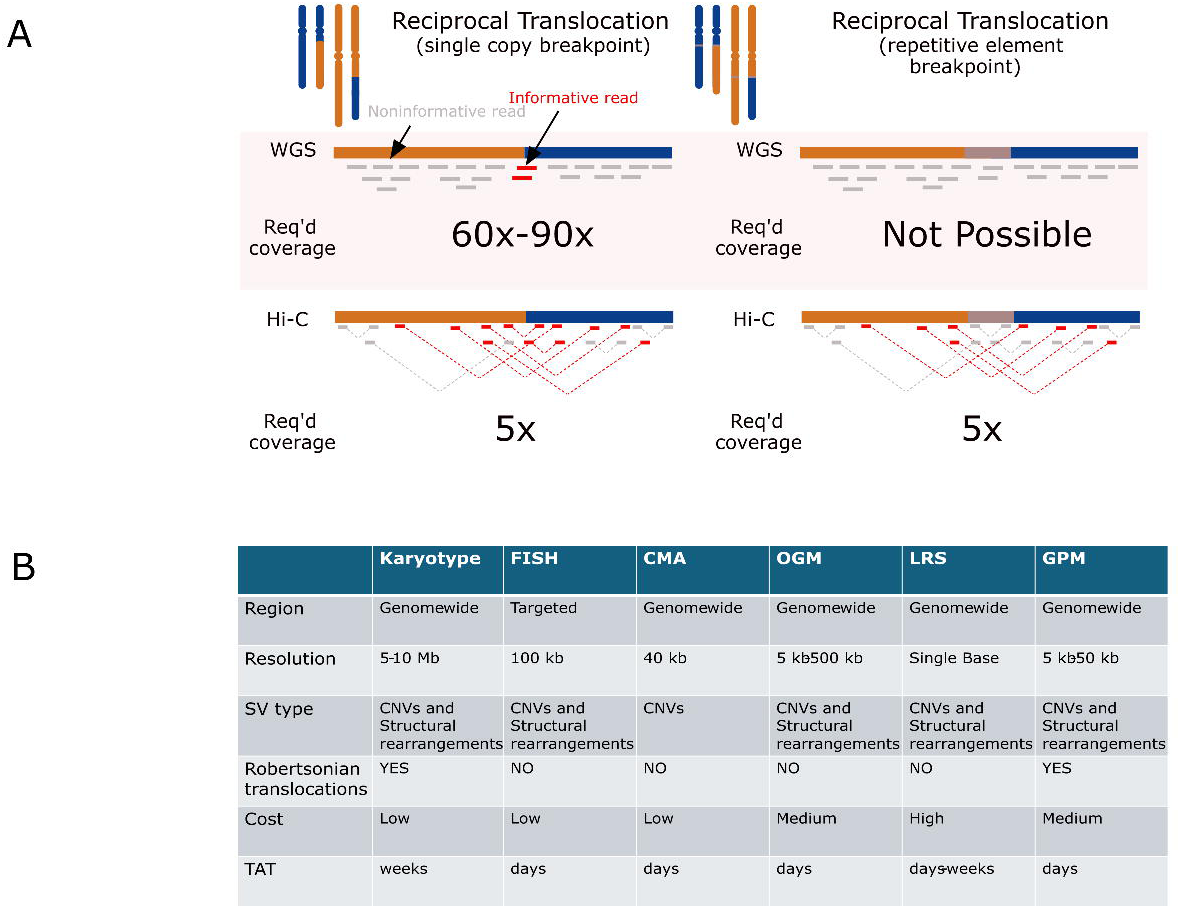
Hi-C enables detection of structural variants at low sequencing coverage in both repetitive and unique regions, compared with other SV-calling methods. (A) Schematic illustrating why Hi-C contact maps reveal rearrangements even at low depth. Two genomic loci are shown in a repetitive (left) and a unique (right) context, corresponding Hi-C contact matrices (bin size 50 kb) display enriched off-diagonal signals (red arrows) at junctions created by a simple inversion event in both contexts, whereas short-read paired-end alignments (blue arcs) are too sparse to call breakpoints reliably. (B) Summary table comparing strengths and limitations of five commonly used SV-detection approaches: short-read WGS, long-read sequencing, optical mapping, Strand-seq, and Hi-C.

A central advantage of GPM is its ability to detect inter-chromosomal translocations and other rearrangements at a lower sequencing coverage than other NGS-based methods, reducing both cost and resource demands without sacrificing sensitivity.. This is largely due to the method’s reliance on spatial chromatin interactions rather than solely on sequence read alignment. For example, novel interactions introduced by translocations between previously unconnected chromosomes or the distinct “bowtie” signatures that result from inversions provide strong, discernible signals in the Hi-C interaction maps (Figure 1A). In our experiments, we were able to identify inter-chromosomal translocations using as little as 2× coverage (Figure 7), which is a significant reduction compared to what traditional WGS would require. Moreover, while intra-chromosomal rearrangements typically require higher coverage due to the more complex background of interactions within a single chromosome, the presence of simultaneous inter-chromosomal changes can effectively lower this threshold. For instance, an intra-chromosomal insertion event is accompanied by a translocation, as we observed in a case involving chromosome 5 (Figure 2D). This structural rearrangement is supported by multiple layers of evidence, including the absence of a stripe in the inter-chromosomal interaction block and the presence of additional interactions in the intra-chromosomal interaction block. These associated signals reinforce the detection of the rearrangement. Essentially, the spatial data allow us to provide additional context to the collinearity of rearrangements, such as breakpoint proximity and orientation, which helps in confidently identifying and characterizing the events even with relatively low sequence coverage.

GPM also excels in detecting complex chromosomal rearrangements that are challenging to identify by conventional sequencing methods. Our analysis revealed that in cases in which traditional cytogenetics identifies more than two chromosomal rearrangements, that these cases tend to harbor additional, cryptic rearrangements that go unnoticed using SOC approaches. In 4 cases with more than two structural rearrangements identified by traditional cytogenetics, we identified additional novel rearrangements in all of them. This observation suggests that such cases may indicate a genomic context that is inherently unstable and prone to further structural disruptions. For example, in one of our cohort cases, chromosomal analysis detected a three-way translocation, yet GPM uncovered additional cryptic alterations—bringing the total to 13 breakpoints distributed across four chromosomes. Notably, a segment on chromosome 7 was found to be dispersed among chromosomes 1q, 4p, and 7p. The spatial cues derived from Hi-C data were crucial here; they enabled us to accurately determine the location, orientation, and extent of each fragment. This level of detail is pivotal, as it not only confirms the rearrangements observed through conventional methods but also refines our understanding of the underlying structural complexity. The ability to map these additional breakpoints with high resolution is critical for understanding the full impact of structural variations on gene regulation and cellular function.

A particularly challenging category of rearrangements involves Robertsonian translocations and whole arm translocations, which typically occur in regions rich in repetitive DNA (Figure 8). Conventional sequencing techniques—including WGS, OGM, and even long-read sequencing—struggle to capture these events due to poor coverage in repetitive regions [22]. In contrast, GPM leverages the three-dimensional spatial organization of chromatin to detect and resolve Robertsonian and whole-arm translocations with a single assay, thereby providing a more complete picture of the genomic landscape (Figure 3).

Beyond structural rearrangements, GPM simultaneously detects CNVs and AOH in a single assay. CNV calls are supported by both shifts in sequencing coverage and disruptions in the Hi-C contact matrix. From our empirical quality-control analyses (Figure 7), we determined that at least 60 million high-quality paired-end reads (∼10× coverage) are required to reliably detect CNVs larger than 25 kb, while AOH regions exceeding 400 kb demand roughly 130 million reads. In silico down-sampling further showed that a 2 Mb deletion present at 40% mosaicism could still be detected with just 30 million reads (∼3×). These limits of detection were evaluated using the cases collected in our study cohort, and in silico dilution and mixture experiments. However, this can highly depend on the sequencing depth, the genomic content of the region and the nature of the events.

GPM also resolves duplication orientation by examining Hi-C heatmap interaction patterns. In human, most duplications proved to be tandem, and we confirmed the two tandem duplications on the X chromosome using Hi-C contact signatures. However, because these patterns arise from relatively short-range contacts, which usually show a higher background signal, deeper sequencing may be necessary for unambiguous orientation calls in some cases. Allelic phasing of multiple CNVs on the same chromosome is another strength—and a challenge—of GPM. By leveraging chromatin contacts that link SNPs across tens to hundreds of kilobases, GPM can assign variants to maternal or paternal haplotypes without parental DNA, in principle. In practice, however, the sparse distribution of informative SNPs and variable contact coverage limit phasing resolution. While current computational methods enable research-grade phasing from GPM data, achieving clinical-grade accuracy will require higher coverage, more robust SNP calling in complex regions, and orthogonal validation.

GPM’s compatibility with archived samples, such as formalin-fixed paraffin-embedded (FFPE) tissues, further underscores its versatility. By capturing preserved chromatin interactions, GPM overcomes the limitations of methods that require high molecular weight or ultra-long DNA (e.g., long-read sequencing or OGM). This robustness is evident in the high pass rate (97%) achieved in our study, even under suboptimal sample conditions.

Hi-C’s ability to visualize the three-dimensional organization of the genome offers a powerful means to link structural variants with functional outcomes. For example, when an inversion occurs, it can physically displace an oncogene away from its upstream enhancers [23–25]. This displacement may either diminish necessary enhancer-mediated activation or inadvertently create new enhancer-promoter contacts that drive aberrant gene activity. Research by Spielmann et al. highlighted that disruptions in topologically associating domains (TADs) due to structural variants can lead to misregulation by altering enhancer-promoter contacts, further underscoring the functional impact of spatial genome organization [26]. These studies collectively indicate that structural rearrangements not only alter the physical layout of the genome but also its regulatory landscape. By disconnecting genes from their native enhancers or juxtaposing them with ectopic regulatory elements, such rearrangements can profoundly affect gene expression and cellular behavior. This integrative perspective—linking structural changes with functional outcomes—provides critical insights into disease mechanisms and points toward potential therapeutic targets.

In current clinical practice, comprehensive SV detection often requires a series of separate tests—karyotyping, FISH, CMA, and sometimes additional follow-up assays—each adding time, cost, and hands-on effort (Figure 8B). GPM replaces this multi-step process with a single Hi-C–based assay that captures balanced and unbalanced rearrangements and CNVs at once. By consolidating what would normally be multiple standard-of-care tests into one streamlined workflow, GPM reduces laboratory burden, cuts turnaround times, and lowers overall costs, while still delivering comprehensive, high-resolution results.

Despite its advantages, GPM is not without limitations. Unlike single-cell assays such as karyotype or FISH, GPM analyzes bulk cell populations, making it challenging to distinguish subclonal abnormalities. Its resolution is dependent on sequencing depth, limiting sensitivity for small-scale variants like single nucleotide variants (SNVs) or indels. Additionally, the computational tools required for Hi-C data analysis may pose adoption challenges for some laboratories. However, these challenges are being addressed by commercial platforms such as the Phase Genomics CytoTerra and are likely to diminish with ongoing advances in technology and bioinformatics.

## Conclusion

In summary, GPM represents a transformative approach to structural variant detection by leveraging spatial genome organization. It offers high sensitivity for complex and mosaic rearrangements, reduced sequencing coverage requirements, and the ability to detect multiple genomic variations—including CNVs and AOH regions—in a single assay. Although challenges remain in resolution and computational analysis, GPM’s robust performance and streamlined workflow position it as a powerful tool for both genomic research and clinical diagnostics.

## Author Contributions

Conceptualization: Y.J.L., S.M.E., H.F.; Funding acquisition: a Brotman Baty Institute grant to H.F., Y.J.L.; NICHD/NIH R44HD104323 to S.M.E.; Writing-original draft: H.F., Y.J.L., S.M.E.; Writing-review & editing: all.

## Ethics Declaration

This project was approved by the institutional review board (IRB) of the University of Washington. Written informed consent was obtained from all participants or their parents.

## Data availability statement

Anonymized data can be made available upon request and with appropriate agreements and human research ethics committee approval.

---

Supplementary Figure 1. A complex chromosomal translocation identified in a balanced carrier. Zoomed-in views of blocks A, B and C reveal breakpoints at gene-level resolution. Block A represents the translocation of a fragment from the short arm of chromosome 4 to chromosome 5, Block B represented the pericentric inversion on chromosome 5, while Block C represents the reciprocal translocation of a fragment from the short arm of chromosome 5 to the short arm of chromosome 4. Hot corner C highlights a pericentric inversion on chromosome 5. This complex rearrangement is also chromosome ideograms at bottom.

Supplementary Figure 2 (A and B) The subtle insertion detected by GPM as part of a complex rearrangements involving chromosome 4 and 5. A represents the absence of a stripe in the heatmap between chromosome 4 and 5, suggesting this fragment is intact on chromosome 5, instead of chromosome 4, in the balanced carrier. B represents the 2-copy state of a DNA fragment on chromosome 5 detected by array in the unbalanced POC sample. These two fragments are of the same size and at the same chromosomal location. (C) Example of a Hi-C heatmap showing an inserted fragment from chromosome 7 (∼719bp) to chromosome 4. (D) The detection of copy gain of the whole short arm of chromosome 4 detected by SNP array in the case with a der(20)t(4;20)(p10;p10).

Supplementary Figure 3. Simulation of the heatmap showing duplications associated with complex rearrangements by GPM.

Supplementary Figure 4. Evaluation of the detection limit of GPM for AOH regions. The CNV with BAF plots illustrate the effects of mosaicism level on the detection of AOH using Hi-C heatmaps. The large AOH region can be detected at mosaicism levels of 90%, 80%, and 60%.

