## Supplementary figures and images for "Evaluation of Genomic Proximity Mapping (GPM) for Detecting Genomic and Chromosomal Structural Variants in Constitutional Disorders"

### Supplementary figure 1

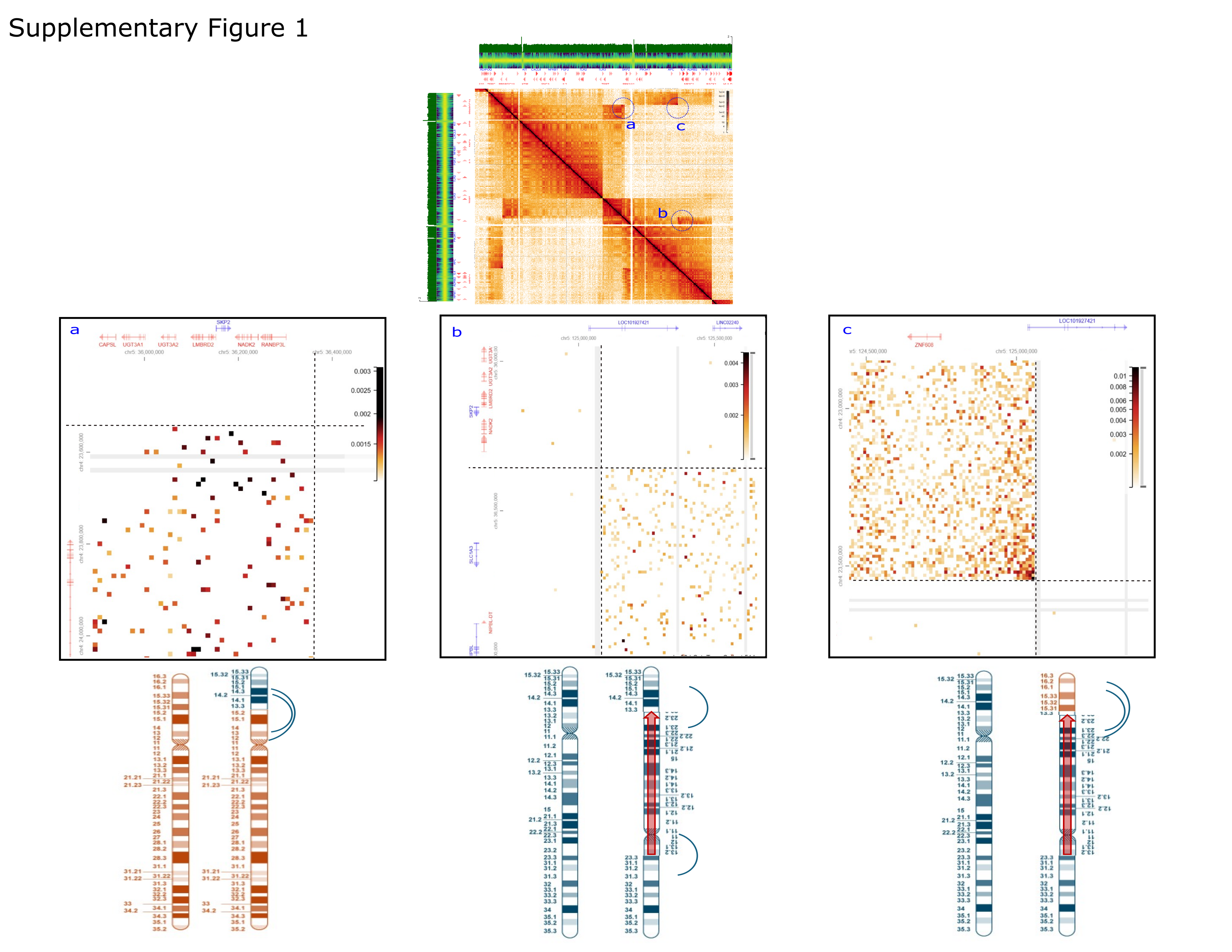

### Supplementary figure 2

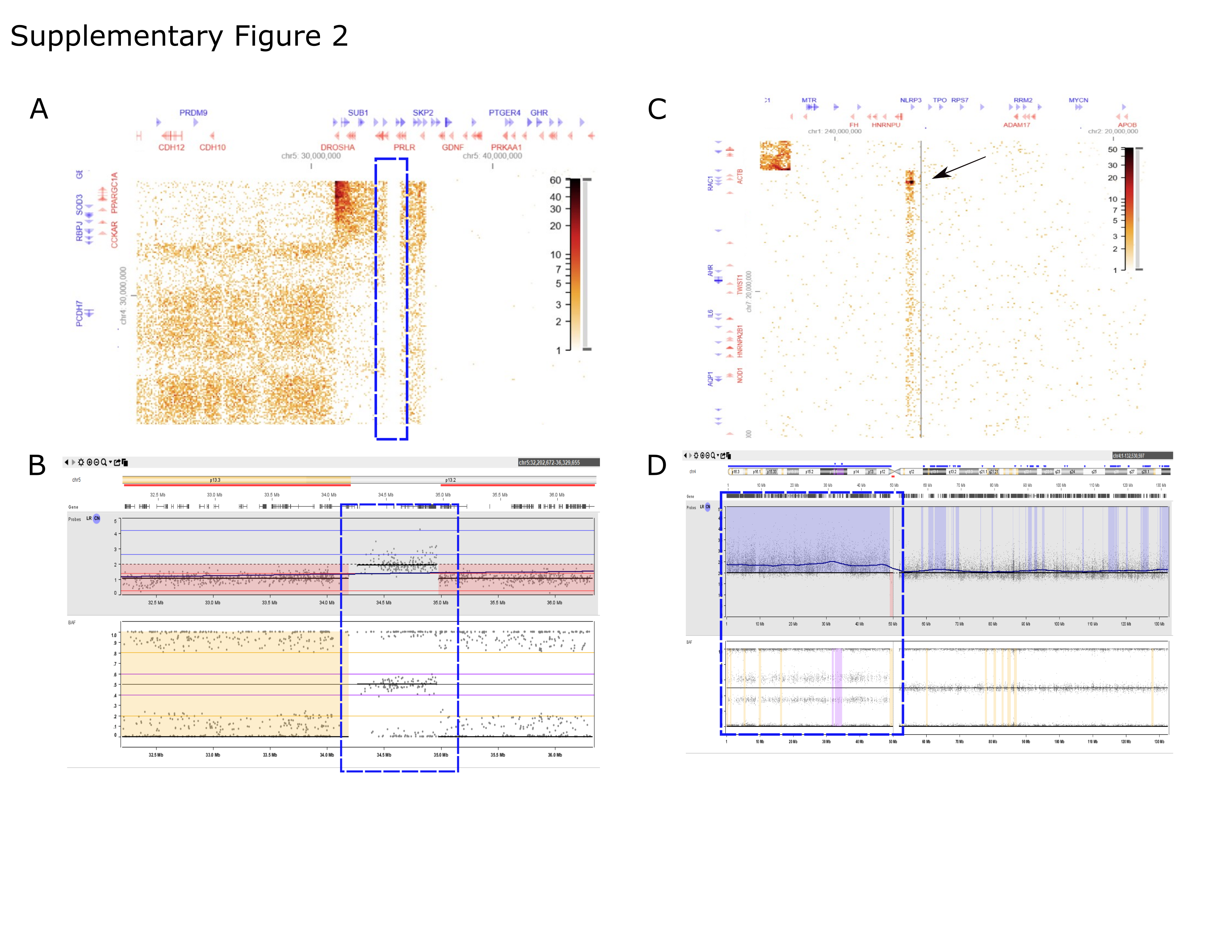

### Supplementary figure 3

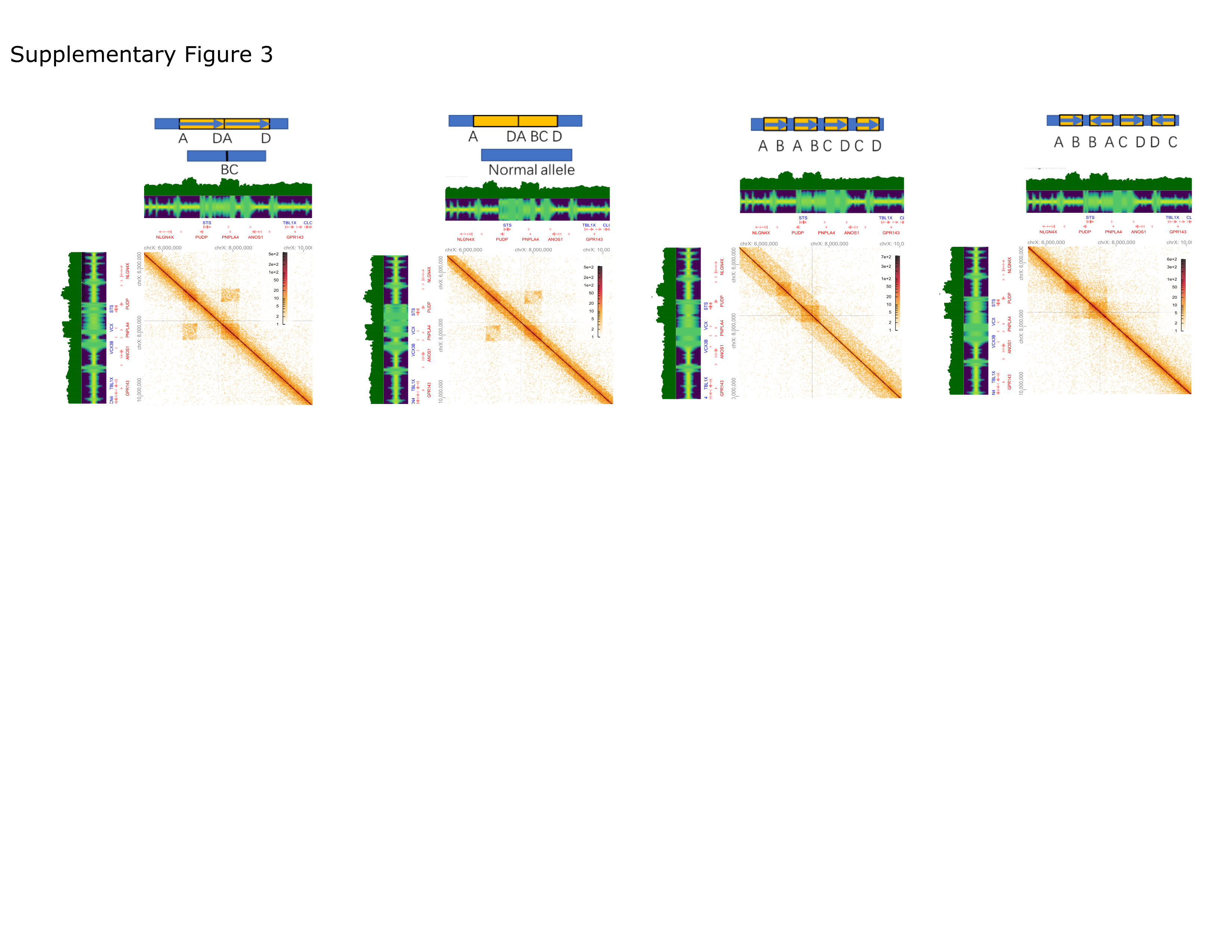

### Supplementary figure 4

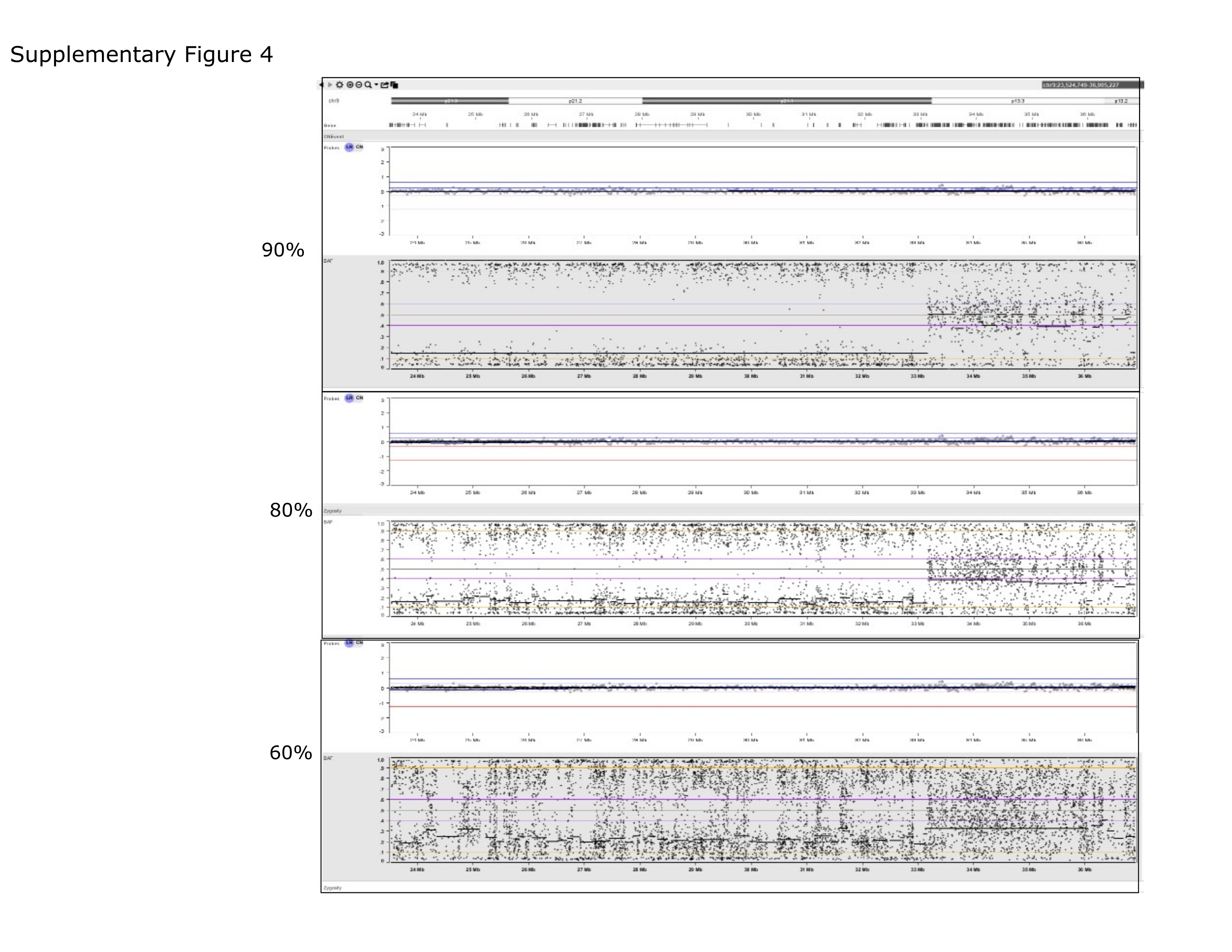
